# Change in Fatigue over 15 months in people the Post-Covid Syndrome

**DOI:** 10.64898/2025.12.20.25342742

**Authors:** Nancy Mayo, Stanley Hum, Lyne Nadeau, Marie-Josée Brouillette, Lesley K Fellows

## Abstract

**Objective:** The objective was to estimate unique patterns of change in fatigue in people with the Post-Covid Syndrome (PCS) over 15 months.

**Design/Subjects:** The Quebec Action for Post-COVID (QAPC) study was a prospective study designed to provide a patient-centered understanding of symptoms, function, and quality of life in a self-identified Quebec sample.

**Methods:** Participants were queried every 3 months about symptoms and function. Fatigue was measured with the 10-item Post-COVID Syndrome Fatigue Severity Measure with a transformed score ranging from 0 no fatigue to 100 extreme fatigue. Group Based Trajectory Analysis (GBTA) was used to identify patterns of longitudinal change.

**Results:** 545 people had an average value of fatigue at baseline of 62.2 / 100 (SD: 21.6); 25% of the cohort had 5 visits. Six trajectories of individual change emerged: two groups with the highest fatigue showed persistence over time; a small group with high fatigue showed improvement; two groups with average fatigue showed improvement over time; and the group with low fatigue showed no emergence of this symptom,

**Conclusion:** High fatigue seems to persist over time while less severe fatigue abates. High fatigue may indicate a sub-syndrome within PCS similar to chronic fatigue syndrome.

**Lay Abstract:** The Post-Covid syndrome (PCS) affected people world-wide and many people still suffer its long term effects. Fatigue is a defining symptom of PCS and there is no evidence-based treatment for this life altering symptom. Using a PCS specific measure of fatigue severity developed using modern measurement theory, six different patterns of longitudinal change were observed. People with the most severe fatigue showed no recovery over time except for a very small group who did recover. People with moderate fatigue showed some improvement. The notion that severe fatigue could be a separate clinical entity needs further study.

## Introduction

The Post-Covid Syndrome (PCS) is defined by symptoms lasting weeks to months after acute infection with SARS-CoV-2. (1–5) In the UK data from the 2021 census estimated that 3% of those infected with SARS-CoV-2 virus had symptoms lasting 4 or more weeks.(2) In Canada by the fall of 2023, 64.4% of adults reported having had COVID-19 (confirmed or suspected) and 19% of infected adults (3.5 million people), had symptoms which persisted for 3 months or more.(6) PCS affected people with all severity of infections(7) from those hospitalized, admitted to intensive care, or staying at home.

Fatigue is the defining symptom of PCS reported in almost all people affected and can be severe, persistent, and debilitating.(8–12)

Despite its prevalence and life-impact, there is little information on how this symptom evolves over time. The most common method used to evaluate evolution of fatigue over time is to query people once at a time distant from the initial COVID-19 event.(13–15) A 2023 systematic review of 12 studies from 11 countries summarizing data from 1,289,044 people(13) reported that 42% of COVID-19 survivors experienced at least one persistent symptom 2 years after COVID-19 and fatigue was the most common affecting 27% of respondents.

A study of health professionals from 21 countries (n=4673) found that 67% reported fatigue as the initial symptom and only half as many 5 months with a small proportion persisting up to one year.(14)

In a 2024 systematic review (15) of 12 studies (n=7,912) with an average follow-up of 2 years, 28% showed persistent fatigue.

A more optimal way of identifying the evolution of fatigue over time is to follow a group of people with PCS longitudinally. For example, Fernández-de-Las-Peñas (16) followed a cohort of 363 people hospitalized for COVID three times over 18 months and reported that 56% of the sample reported fatigue at 6 months with 12% resolving by 12 months. For those with fatigue beyond 12 months, 15% resolved by 18 months. A small proportion of people (7%) initially without fatigue at 6 months reported emergence by 12 months. A portion (5%) of those resolving rebounded. The RECoVERED cohort(17) comprised 303 people recovering from COVID-19 who were followed over 6 time points up to12 months. For those with at least one assessment of fatigue, measured using the Short Fatigue Questionnaire (range 4–28), prevalence depended on severity of initial infection with a range of 17% among those with mild infection to 45% among those with severe infection. Recovery of fatigue severity over time was estimated at −0.35 units (95% CI: –0.45 to –0.25) per month with little recovery beyond 6 months.

The study reported here contributes to the knowledge base on evolution of fatigue using a different statistical approach based on identifying individual patterns of longitudinal change.(18) The objective was to estimate, in a sample of people self-identifying with PCS, unique patterns of change in fatigue over 15 months and characteristics of people with different longitudinal trajectories.

## Methods

The data for this longitudinal study came from the Quebec Action for Post-COVID (QAPC) study designed to provide a patient-centered understanding of symptoms, function, and quality of life in a self-identified Quebec sample. A description of the sample and results have been described in detail in a previous study.(19) This study reports on the evolution of fatigue in the full cohort recruited from September 23, 2022, until September 15, 2023 and followed for up to 15 months. The project (2022-8066) was approved by the Research Ethics Board of the McGill University Health Centre. People interested in participating were directed to the QAPC website to register.

### Population

Residents of Quebec age 18 and over were eligible if they currently had symptoms occurring 4 or more weeks post onset of symptoms of the COVID-19 infection, with or without a positive test. The sample was assembled from multiple sources: most participants were reached through French-and English-language media (radio) and social media, with some contacted via email outreach to a waitlist for a post-COVID research clinic. Recruitment was through the QAPC website. The study coordinator recorded their contact information, generated a study identification number and invited them into the study. Upon invitation, they were directed to an online web-portal “Research Electronic Data Capture” (REDCap) to enter their unique identification number, allowing them entry into the data capture platform. Following an e-consent process, they recorded their health outcomes information. All were asked to consent to open data sharing for secondary analyses and to be re-contacted for additional studies.

### Measurement

Participants were queried every 3 months. The complete portfolio of measures for this study have been described in detail in the previous publication.(19) This current analysis focuses on fatigue measured with the Post-COVID Syndrome Fatigue Severity Measure. Rasch analysis was used to identify those fatigue-related items included on the QAPC platform specifically to reflect the fatigue construct that fit linear continuum to form a measure with mathematical properties. The methods for developing the PCS Fatigue Severity Measure have been reported previously.(20) Briefly, 10-items fit the Rasch model. The items and response options are given in Table I. The scoring range is 0 to 21 with 21 indicating the highest fatigue; for ease of interpretation, this score was transformed to be out of 100. There was evidence for concurrent validity and important differences across groups of people with different effects of PCS. Correlations with converging constructs of physical, social and cognitive function and measures of health aspects of quality of life were >0.5. Evidence for an important difference was provided based on the difference between people unable to work because of PCS and those still working. On a scale of 0 to 20, this value was 5.1 (effect size: 1.23); scaled to be out of 100, this value would be 25. Using an medium effect size of ½ SD is a more reasonable minimal important difference.(21)

**Table I.** Items Fitting the Post-COVID Syndrome (PCS) Fatigue Severity Measure.

| Item |
| --- |
| 1. How much time during the past 4 weeks did you have a lot of energy?<br>*All of the time [0]; Most of the time [1]; Good bit of the time [2]; Some of the time [3]; A little or none of the time [4]* |
| 2. How much time during the past 4 weeks did you feel worn out?<br>*None or a little of the time [0]; Some of the time [1]; Good bit, most, or all of the time [2]* |
| 3. Throughout the past month how much have you experienced dead, heavy feeling after starting to exercise<br>*Not present [0]; Mild/moderate [1]; Severe/very severe [2]* |
| 4. Throughout the past month how much have you experienced next day soreness or fatigue after non-strenuous, everyday activities<br>*Not present [0]; Mild/moderate [1]; Severe/very severe [2]* |
| 5. Throughout the past month how much have you experienced being mentally tired after the slightest effort<br>*Not present [0]; Mild/moderate [1]; Severe/very severe [2]* |
| 6. Throughout the past month how much have you experienced being physically drained or sick after mild activity<br>*Not present [0]; Mild/moderate [1]; Severe/very severe [2]* |
| 7. Do you have to stop for breath after walking about 100 meters or after a few minutes on level ground?<br>*No [0]; Yes [1]* |
| 8. Are you too breathless to leave the house, or breathless when undressing?<br>*No [0]; Yes [1]* |
| 9. How would you describe your fatigue in the past week?<br>*Most often I never felt so tired that I had to rest [0]; I felt so tired that I had to rest one or more times throughout the day [1]; I felt so tired that I had to rest most of the day [2]* |
| 10. On a scale from 0 to 10 where 0 is no fatigue and 10 is the worst fatigue imaginable, how would you rate your usual degree of fatigue?<br>*0-4 [0]; 5-8 [1]; 9-10 [2]* |
| Note: Raw scores range from 0 to 21 with higher scores indicating more fatigue and then rescored to range from 0 to 100 for ease of interpretation and presentation of results. |

### Statistical Methods

The cohort was described on variables related to the fatigue construct. The numbers of people with different patterns of follow-up are presented as well as the numbers of people with 1 to 6 visits. Baseline fatigue values were compared across groups with different numbers of follow-up visits using linear regression. Average changes over time were estimated for the whole cohort using a linear mixed model with time as the random effect and using an unstructured covariance matrix.

The main analysis was Group Based Trajectory Analysis (GBTA) (22) which is a semi-parametric method of grouping together people with similar longitudinal trajectories of fatigue severity. Only people with 2 or more data points were included in the analysis. Parameters related to fit of the GBTA model and describing the trajectories are Akaike information criterion (AIC) and Bayesian Information Criteria (BIC), posterior probability of trajectory group assignment, numbers of people theoretically assigned to trajectories, numbers of people assigned to trajectories based on highest probability of group membership, intercept, degree of linear effect, degree of quadratic effect.

Models are built up from one to n trajectories. First all trajectories are described using both a linear and quadratic term and these are dropped if they do not achieve statistical or clinical relevance. The best fitting model is based on minimizing AIC and BIC while considering existing knowledge in the field. Fit to the model is also demonstrated if the posterior probabilities of trajectory membership is >70%.

Regression models were used to identify whether trajectory membership was related to age (linear regression), sex (logistic regression), and time since last COVID episode (quantile regression at median).

### Sample size

The QAPC cohort was assembled to describe symptom patterns longitudinally so no *a priori* estimates of sample size were made. There are no closed formulae for calculating sample size for GBTA. The greater the sample size, the greater are the number of trajectories that can be detected but sample sizes are typically in the hundreds.(23)

## Results

Table II presents the characteristics of the cohort participants (mean age: 48.8 years; 76.1% women) at their first assessment. The time from last COVID episode was 251 days and more for half of the sample and 28.7% had COVID more than once. Other information is presented on factors that are likely to be related to fatigue. People had a range of roles and responsibilities, and half had worries about money.

**Table II.** Characteristics of the QAPC Sample (n=555) at First Assessment.

| **Variable** | **N<sup>a</sup> (%) Mean (SD) [Min-Max]** |
| --- | --- |
| Sex: Men/Women/Other | 121/407/7 (22.6%/76.1%/1.3%) |
| Age (mean) | 48.8 (12.2) [18-83] |
| Time since first COVID: Median (25<sup>th</sup>-75<sup>th</sup>) | 287 (168-248) |
| Time since last COVID: Median (25<sup>th</sup>-75<sup>th</sup>) | 251 (129-251) |
| COVID more than once | 151 (28.7%) |
| Roles and responsibilities outside work |  |
| Pre-school children | 49 (8.9%) |
| School-age children | 187 (33.8) |
| Caring for pets | 283 (51.1%) |
| Shopping | 422 (76.3%) |
| Meal preparation | 425 (76.9%) |
| Household management | 444 (80.9%) |
| Money to meet needs: not completely | 265 (49.7%) |
| Worry about money | 308 (57.8%) |
| Post-exertional malaise (PEM) | 458 (83.7%) |
| Need to rest during the day |  |
| No | 50 (9.1%) |
| 1+ times during the day | 297 (54.3%) |
| Most of the day | 200 (36.6%) |
| PTSD ($\geq 3/5$ symptoms) | 131 (25.1) |
| Breathlessness |  |
| SOB walking 100 meters on level ground | 89 (16.3%) |
| SOB dressing / can't leave house | 32 (5.8%) |
| Prescribed medication for PCS symptoms | 200 (37.5%) |
| Lonely sometimes or often [10.4%] | 325 (61.0%) |
| Irritable often to always | 112 (21.0%) |

<sup>a</sup>Numbers of participants with missing data on variables ranges from 0 to 31.

Table III describes the cohort in terms of fatigue, other symptoms, function, quality of life. Normative data is provided when available. At this first assessment, PCS fatigue averaged 62.2 out of a maximum fatigue value of 100. The burden of other symptoms was high considering that values of 40 or more out of 100 reach the clinically concerning level. For function, values for the QAPC were consistently lower than age-expected norms.

**Table III.**
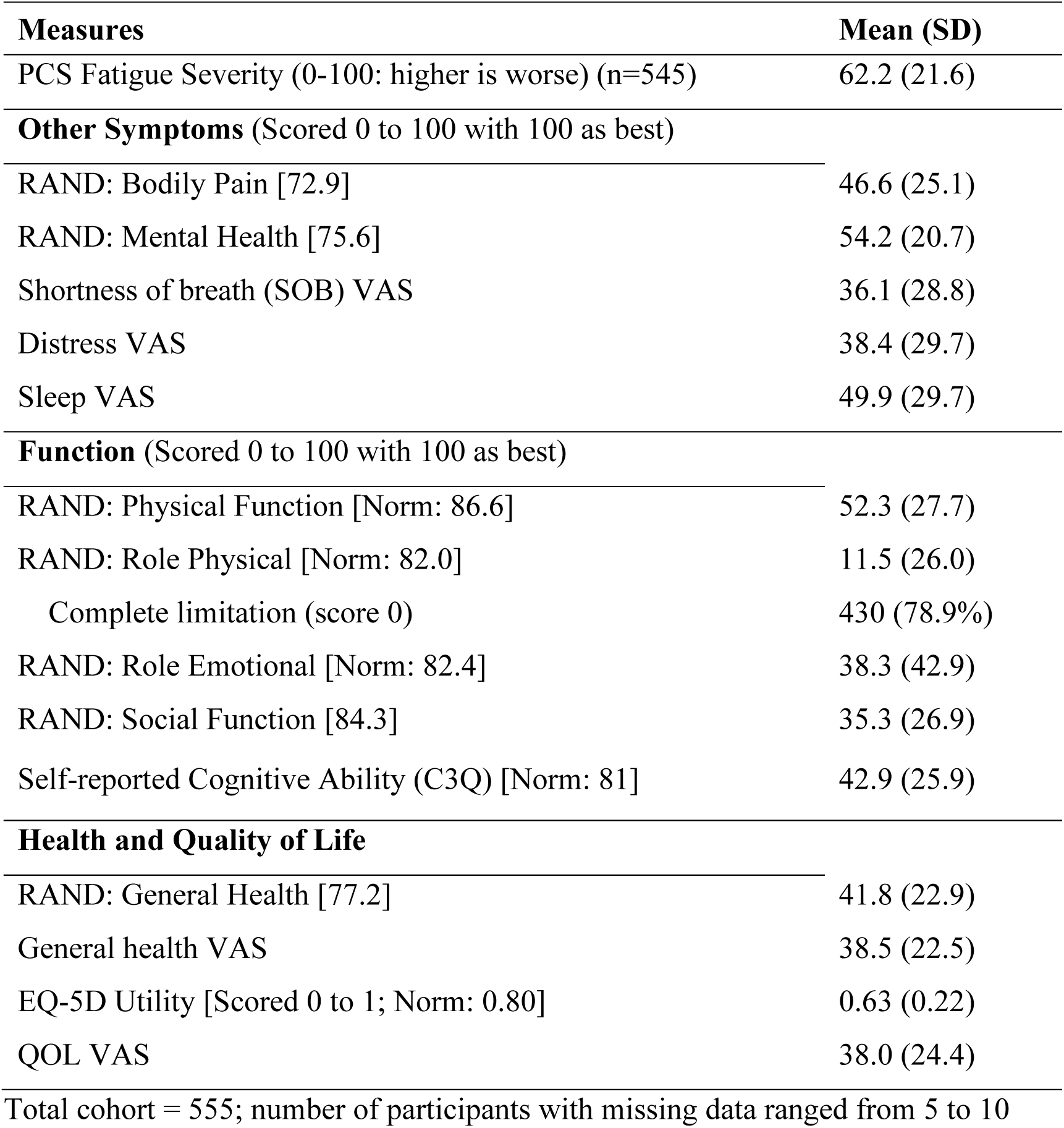
Values on Measures of Symptoms, Function, Health, and QOL Outcomes.

| **Measures** | **Mean (SD)** |
| --- | --- |
| PCS Fatigue Severity (0-100: higher is worse) (n=545) | 62.2 (21.6) |
| **Other Symptoms** (Scored 0 to 100 with 100 as best) |  |
| RAND: Bodily Pain [72.9] | 46.6 (25.1) |
| RAND: Mental Health [75.6] | 54.2 (20.7) |
| Shortness of breath (SOB) VAS | 36.1 (28.8) |
| Distress VAS | 38.4 (29.7) |
| Sleep VAS | 49.9 (29.7) |
| **Function** (Scored 0 to 100 with 100 as best) |  |
| RAND: Physical Function [Norm: 86.6] | 52.3 (27.7) |
| RAND: Role Physical [Norm: 82.0] | 11.5 (26.0) |
| Complete limitation (score 0) | 430 (78.9%) |
| RAND: Role Emotional [Norm: 82.4] | 38.3 (42.9) |
| RAND: Social Function [84.3] | 35.3 (26.9) |
| Self-reported Cognitive Ability (C3Q) [Norm: 81] | 42.9 (25.9) |
| **Health and Quality of Life** |  |
| RAND: General Health [77.2] | 41.8 (22.9) |
| General health VAS | 38.5 (22.5) |
| EQ-5D Utility [Scored 0 to 1; Norm: 0.80] | 0.63 (0.22) |
| QOL VAS | 38.0 (24.4) |

Total cohort = 555; number of participants with missing data ranged from 5 to 10

Table IV presents average values on the PCS Fatigue Severity measure according to pattern of follow-up assessments. The average values of people with 1, 2, 3, 4, 5 and 6 assessments showed not pattern indicative of response bias. In comparison to people with only 1 assessment (PCS Fatigue Severity: 61.0), only the group with 4 assessments showed a statistically and meaningful difference in fatigue severity (68.2). Also presented are average fatigue values for each of these groups defined by follow-up pattern and there is very little change over time.

**Table IV.** Mean (SD) Fatigue Scores by Visit According to Pattern of Visits and Overall Patterns.

| By Pattern<sup>a</sup> | Visit | N with data / N | Visit 1 Mean | Subsequent Visit |
| --- | --- | --- | --- | --- |
| 1-0-0-0-0-0-0 | 1 | 177/182 | 61.0 (23.2) | Referent group |
| 1-1-0-0-0-0-0 | 1 | 89/90 | 62.8 (21.6) |  |
|  | 2 | 63/90 |  | 58.7 (23.3) |
| 1-1-1-0-0-0-0 | 1 | 50/50 | 65.1 (19.2) |  |
|  | 2 | 41/50 |  | 59.9 (21.2) |
|  | 3 | 34/50 |  | 54.1 (23.6) |
| 1-1-1-1-0-0-0 | 1 | 67/68 | 68.2 (20.4) | p=0.0194 vs. referent |
|  | 2 | 61/68 |  | 63.8 (21.6) |
|  | 3 | 61/68 |  | 64.0 (22.4) |
|  | 4 | 57/68 |  | 65.2 (21.8) |
| 1-1-1-1-1-0-0 | 1 | 62/62 | 56.8 (20.9) |  |
|  | 2 | 55/62 |  | 56.4 (19.9) |
|  | 3 | 52/62 |  | 55.5 (21.5) |
|  | 4 | 51/62 |  | 50.8 (21.0) |
|  | 5 | 46/62 |  | 51.5 (22.4) |
| 1-1-1-1-1-1-0 | 1 | 81/84 | 60.0 (20.8) |  |
|  | 2 | 74/84 |  | 59.0 (20.8) |
|  | 3 | 81/84 |  | 56.2 (21.8) |
|  | 4 | 79/84 |  | 56.7 (21.6) |
|  | 5 | 74/84 |  | 55.2 (22.0) |
|  | 6 | 73/84 |  | 54.6 (22.5) |
| 1-1-1-1-1-1-1 | 1 | 19/19 | 67.7 (19.2) |  |
|  | 2 | 15/19 |  | 60.9 (18.4) |
|  | 3 | 19/19 |  | 58.9 (19.4) |
|  | 4 | 17/19 |  | 56.7 (22.7) |
|  | 5 | 18/19 |  | 55.8 (21.9) |
|  | 6 | 19/19 |  | 59.8 (20.8) |
|  | 7 | 18/19 |  | 59.6 (20.3) |
| By Visit | 1 | 545 | 62.2 (21.6) |  |
|  | 2 | 309 | 59.6 (21.3) |  |
|  | 3 | 247 | 57.9 (22.1) |  |
|  | 4 | 204 | 57.6 (22.1) |  |
|  | 5 | 138 | 54.0 (22.0) |  |
|  | 6 | 92 | 55.7 (22.2) |  |
|  | 7 | 18 | 59.6 (20.3) |  |
| Effect of time<sup>b</sup> | | $\beta=-0.44$ (SE: 0.08; $p<0.0001$) | | |

<sup>a</sup>pattern indicates how many visits were completed.
<sup>b</sup>time in months; adjusted for age

Average values across assessments also show little differences from the whole sample at baseline (mean 61.0). The results of the linear mixed effects model provide an estimate of change of time measured in months. The estimated change over time was −0.44 units (SE: 0.08; 95% CI: −0.28 to −0.60) which was highly significant but not clinically important as over a 15-month period this would translate to an average change of 6.6 units (out of 100) which is the equivalent to an effect size of 0.31 (6.6/SD of 21.6).

As the average values presented in Table IV do not depict longitudinal change at the individual level, GBTA was used and the results of the best fitting model are shown in Figure 1 and key parameters related to the trajectories given in Table V. Figure I shows that 6 trajectories of individual change emerged from the data. Two groups, (labeled 6 and 5) entered the cohort with higher-than-average fatigue and were predicted to remain with this high level over 15 months. In Table V, these two groups are labeled “Very high, persistent” and “High, persistent”. A small group, labeled 4 in Figure 1, entered the cohort with high fatigue, similar to group 5, but rapidly improved. In Table V, this group is labeled “High fatigue, resolving”. Group 3, entered with average fatigue and showed improvement as did group 2. In Table V, these groups were labeled “Medium-high, improving” and Medium, improving”. The 6^th^ group (“Low, stable”) entered with low fatigue remained low over time.

**Figure 1.**
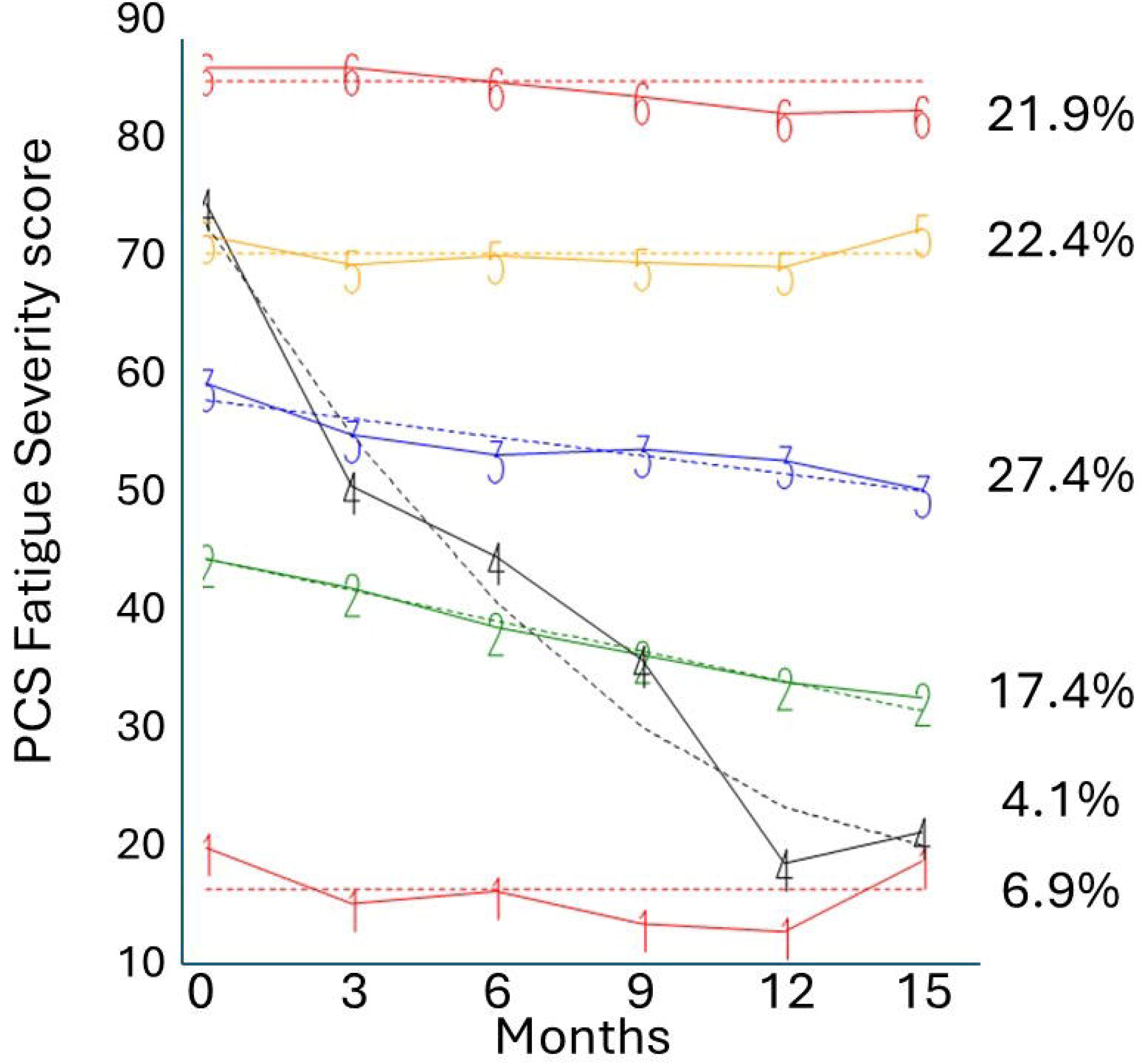
Trajectories of Post-COVID Fatigue Severity.

**Table V.**
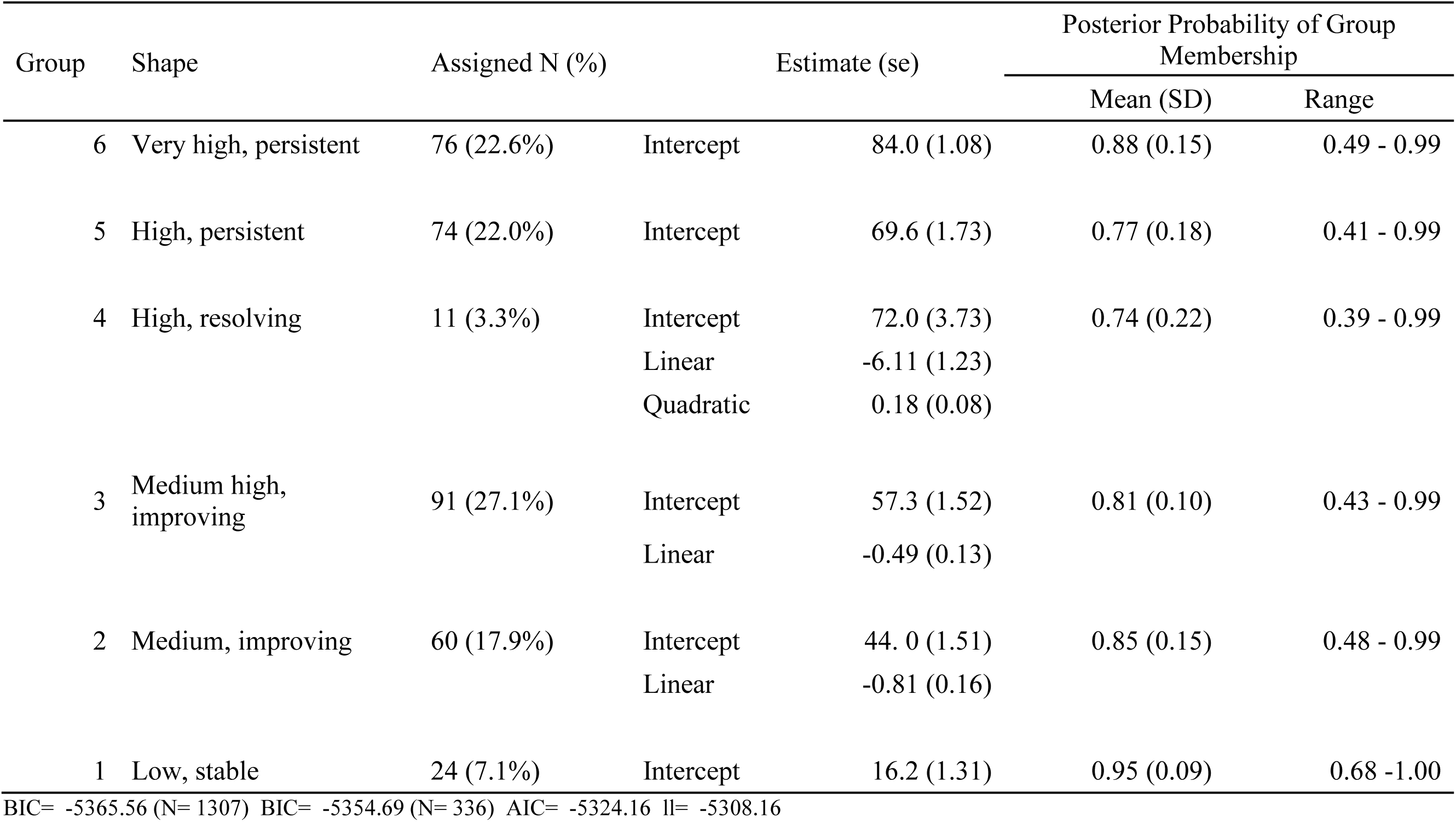
Characteristics of the Fatigue Trajectories of 336 People with at Least 2 Measurements.

| Group | Shape | Assigned N (%) | Estimate (se) | Posterior Probability of Group Membership |  |
| --- | --- | --- | --- | --- | --- |
|  |  |  |  | Mean (SD) | Range |
| 6 | Very high, persistent | 76 (22.6%) | Intercept<br>84.0 (1.08) | 0.88 (0.15) | 0.49 - 0.99 |
| 5 | High, persistent | 74 (22.0%) | Intercept<br>69.6 (1.73) | 0.77 (0.18) | 0.41 - 0.99 |
| 4 | High, resolving | 11 (3.3%) | Intercept<br>72.0 (3.73)<br>Linear<br>-6.11 (1.23)<br>Quadratic<br>0.18 (0.08) | 0.74 (0.22) | 0.39 - 0.99 |
| 3 | Medium high, improving | 91 (27.1%) | Intercept<br>57.3 (1.52)<br>Linear<br>-0.49 (0.13) | 0.81 (0.10) | 0.43 - 0.99 |
| 2 | Medium, improving | 60 (17.9%) | Intercept<br>44.0 (1.51)<br>Linear<br>-0.81 (0.16) | 0.85 (0.15) | 0.48 - 0.99 |
| 1 | Low, stable | 24 (7.1%) | Intercept<br>16.2 (1.31) | 0.95 (0.09) | 0.68 - 1.00 |

BIC= -5365.56 (N= 1307) BIC= -5354.69 (N= 336) AIC= -5324.16 ll= -5308.16

Table V also provides the estimated cohort entry values for these trajectories and whether there were linear and quadratic terms. All trajectories had average posterior probabilities greater than 70%. The estimated change over time for the two highest groups was 0 (groups 6 and 5); change for group 4 was −6.11 moderated by a quadratic term of +0.18. The two groups with medium fatigue (groups 3 and 2) improved over time by −0.49 and −0.80 units per month; the group with the least fatigue also showed 0 change. The change value estimated for the cohort as a whole was −0.44 units (provided in Table IV) closest to the decline only for group 3, comprising 91 people.

The information presented in Table VI shows that sex and time since the last COVID episode did not relate to trajectory membership but that people in the two best trajectories of fatigue were older than the other groups. For these comparisons, the sample of people who did not have any follow-up assessments was the referent group.

**Table VI.** Characteristics of Participants Assigned to Trajectories of PCS Fatigue.

| Group | Shape | Assigned N (%) | Age Mean (SD) | % Women | Time since last COVID event (days) Median |
| --- | --- | --- | --- | --- | --- |
| 6 | Very high, persistent | 76 (22.6%) | 47.3 (13.5) | 84.1% | 269 |
| 5 | High, persistent | 74 (22.0%) | 47.9 10.0() | 83.1% | 244 |
| 4 | High, resolving | 11 (3.3%) | 47.8 (12.1) | 90.9% | 220 |
| 3 | Medium high, improving | 91 (27.1%) | 48.5 (12.7) | 79.1% | 229 |
| 2 | Medium, improving | 60 (17.9%) | 56.0* (11.6) | 73.7% | 192 |
| 1 | Low, stable | 24 (7.1%) | 53.3* (13.5) | 75.0% | 239 |
| NA | Referent | 219 | 47.3 (12.9) | 76.1% | 253 |

NA: only one time point; referent category
\* statistically different from referent

## Discussion

This study found that people with a high degree of fatigue were estimated to remain at this high level over the duration of the study except for a small group of people who showed a resolution (see Figure I). People with an average degree of fatigue showed some improvement over time and those with lower-than-average fatigue remained low. Unlike the study by Fernández-de-Las-Peñas et al.,(16) we did not identify a group that showed an increase in fatigue or bounce back over time. In our study, there were no differences in people in the different fatigue trajectories on age, sex, or time since last COVID events indicating that other reasons are implicated.

The advantage of using GBTA in this study was that variability in degree of change over time can be visualized. Average change over the whole sample was also estimated and reported in Table IV as −0.44 unit per month (SE: 0.08; p<0.0001) which, over 15 months, translates to an decrease in fatigue severity of 6.6 points when related to the SD of 21.6 yields an effect size of 0.31.

This average degree of change was supported by the data in Table IV showing average values over time. However, only one group showed a similar degree of improvement, group 3 (n=91; 27%) with an estimated change of −0.49 unit (SE: 0.13). In a previous study from the Netherlands following 303 people for one year, the average change on a fatigue measure with a scoring range from 4 to 28 was −0.35 units per month (95% CI: −0.25 to −0.45), approximately 1.4 units on a 0 to 100 scale.(24)

The results of this study showed no resolution of the most severe fatigue over 15 months and this pattern is similar to the course of other health conditions like Myalgic Encephalomyelitis/ Chronic Fatigue Syndrome (ME/CFS).(25) Given the large numbers of people with PCS, investigating similarities between people with PCS and ME/CFS could prove insights for both conditions.

It was noteworthy that 37% of participants reported being prescribed one or more medications for symptoms of PCS (see Table II). Michael et al.(26), in earlier work on the QAPC cohort, found that the median number of prescribed medications was 2 and, for 11.7% of the cohort prescribed medications, the pattern met criteria for polypharmacy with the potential risk for clinically significant drug-drug interactions,. Drugs targeting the nervous system were predominant at 54.5% and many of these drugs would increase fatigue and brain fog. This study also found that the severity of fatigue was equally high in those prescribed or not medications for PCS symptom management. This emphasizes the need to understand fatigue defining illnesses to avoid inappropriate prescribing.

This study has the limitations associated with all self-identified cohorts. There is no base population to identify selection biases. Studies based on hospitalized participants do not represent the population with COVID. In most parts of the world, only a small fraction of those with COVID infection were hospitalized. In the USA, this is estimated at 1 to 2%;(27, 28); in Canada, this proportion was approximately 6% early in the pandemic and 1% as vaccination coverage increased.(6)

People most concerned are most likely to respond and so estimates of impact are over estimated. People who drop out may drop out for reasons related to fatigue, they may have improved and returned to normal life, or they may have deteriorated and no longer motivated to respond. Information presented in Table IV showed that baseline level of fatigue did not differ by pattern of response. GBTA also has the advantage of assigning people to trajectories based on the data they have and the posterior probabilities provide an indication of the confidence there is in this trajectory with 70% considered to demonstrate good fit.

## Conclusion

This study on the trajectory of fatigue among people with PCS supports the hypothesis that a high degree of fatigue likely indicates a different pathological process than that which underlies less severe fatigue and could inform the assessment and management of this distressing symptom for people with other fatigue defining illnesses.

## Data Availability

All data produced in the present study are available upon reasonable request to the authors

## Acknowledgements

None

